# Epidemiological Analysis of Confirmed Mpox Cases in Burundi, July to September 2024

**DOI:** 10.1101/2024.09.27.24314511

**Authors:** Alexis Nizigiyimana, Francois Ndikumwenayo, Sarah Houben, Martin Manirakiza, Monique van Lettow, Laurens Liesenborghs, Placide Mbala-Kingebeni, Anne W. Rimoin, Isaac I. Bogoch, Jason Kindachuk

## Abstract

This study analyzes mpox cases in Burundi from July to September 2024, following the introduction of Clade Ib. We analyzed 607 samples from the whole population of suspected cases in the study period where 25.4% of the samples tested positive via PCR. Children under 15 comprised 55.2% of cases, with a higher proportion of female children testing positive. Geographic analysis demonstrates case concentration in Bujumbura Mairie (59.1%). These findings highlight the importance of age- and sex-specific interventions for outbreak containment and the need for targeted public health strategies in Burundi.

## INTRODUCTION

Mpox is a zoonotic viral infectious disease first identified in the Democratic Republic of the Congo (DRC) in 1970 (1-3). Monkeypox virus (MPXV) historically circulated in the tropical forest regions of Central and West Africa, where it is considered endemic (4, 5). The virus is subclassified into two clades: clade I, formerly known as the Congo Basin (Central Africa) clade, and clade II, formerly known as the West African clade. Clade II is further subdivided into two subclades: IIa and IIb, with the latter responsible for the global epidemic in 2022 (6, 7).

Historically, zoonosis has been the primary driver of MPXV infections in humans with secondary infections through human-to-human contact. However, Clade IIb MPXV infections during the 2022 global epidemic were linked primarily to sustained human-to-human transmission (8) including among men who have sex with men (9). The emergence of Clade Ib MPXV has demonstrated similar transmission characteristics including linkage to sexual networks and sustained human-to-human transmission (10). However, Clade Ib mpox cases have not been overrepresented among males. Further, recent expansion of Clade Ib into North Kivu, DRC, has included probable transmission through non-sexual or intimate contacts including children.

While there may be reporting biases complicating a more wholesome understanding of mpox epidemiology, containment and mitigation strategies in this ongoing public health emergency must also consider the context-dependencies of these differences.

From January 2023 to present, broad geographic expansion of Clade I MPXV in the DRC has resulted in the largest mpox outbreak in recorded history among all endemic countries.

Concerningly, this national outbreak has included the expansion of MPXV to all provinces in DRC as well as the emergence of a new virus subclade, Clade Ib, associated with sustained human-to-human transmission (10). While first reported in South Kivu province, DRC, Clade Ib has undergone rapid regional expansion that has included introduction to North Kivu, DRC (11), as well as dissemination to neighbouring countries in East Africa, including Uganda, Rwanda, Kenya, and most notably Burundi.

Here, we provide an analysis of demographic and epidemiological considerations for mpox in Burundi using mpox case data from July to September 2024.

## RESULTS

### Mpox case demographics in Burundi

We analyzed national mpox testing and demographic data acquired in Burundi from 03 July 2024 to 09 September 2024 (week 27 to week 37). During this period, a total of 607 samples were tested by PCR for MPXV (Figure 1). Of these, men comprised 305 samples (50.2%) and women comprised 302 samples (49.8%). For all samples, 154 samples (25.4%) tested positive, 448 samples (73.8%) tested negative, and 5 samples (0.8%) were indeterminate. Of the positive samples, males comprised 51.9% of cases (80/154) and women comprised 48.1% (74/154).

**Figure 1:**
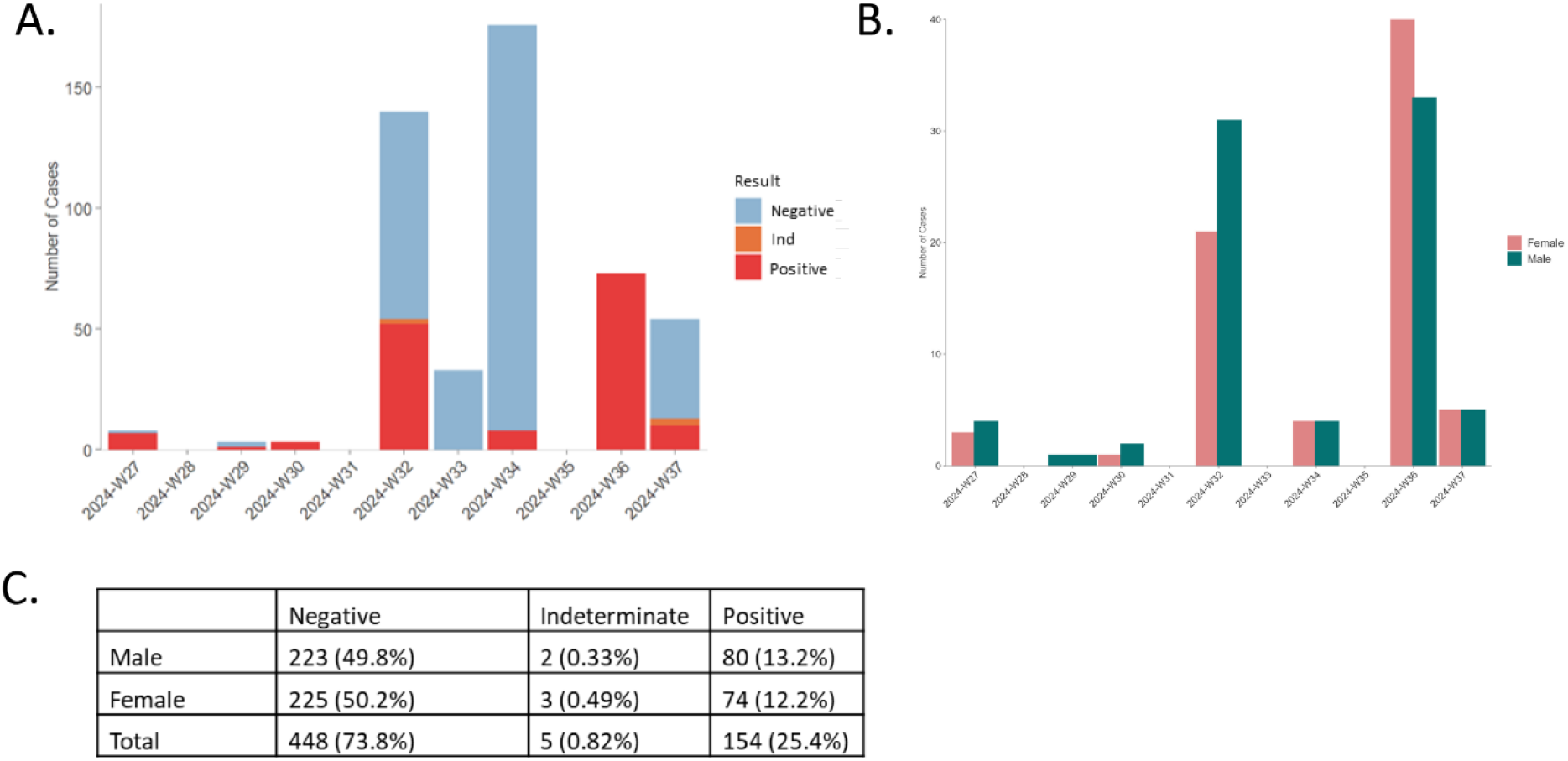
Demographics of mpox test results in Burundi from week 27 to week 37. A) mpox cases by result and week of reporting; B) mpox cases by sex during reporting period; C) mpox testing results by sex during reporting period.

In analyzing age demographics, the median age of all individuals tested was 7 years (IQR: 2-18 years) with an age range of <6 months to 72 years (Figure 2). For mpox-positive cases, the median age was 9.5 years (IQR: 3-25 years). The median age of positive cases was significantly higher for males as compared to females at 17.5 years and 6 years, respectively (p<0.018). An age pyramid for positive mpox cases separated by sex is presented in Figure 2C. Overall, children <15 years comprised the majority of all positive cases (85/154; 55.2%) with cases highest among those 0-4 years followed by those 5-9 years (Figure 2C).

**Figure 2:**
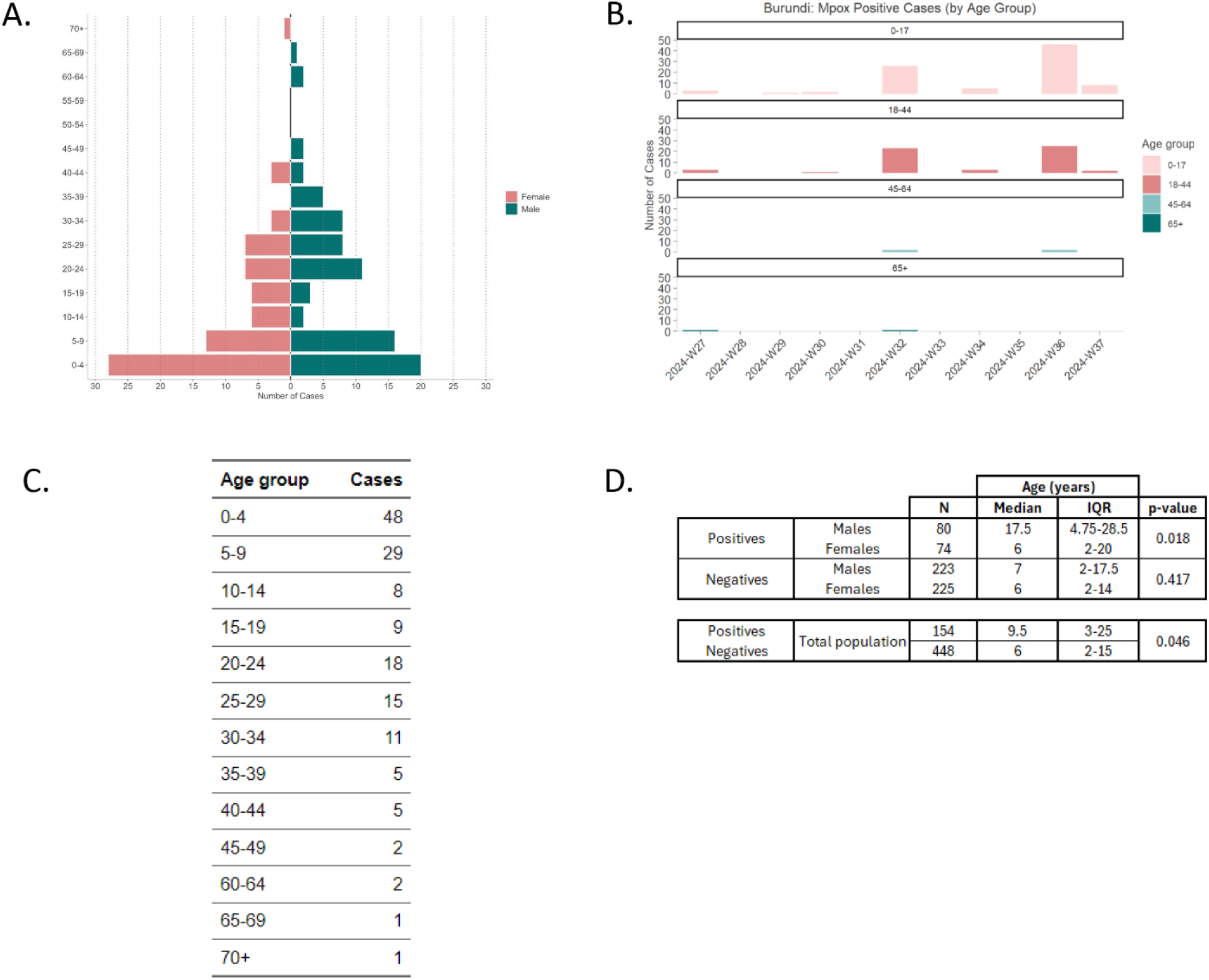
Age demographics for individuals tested for mpox in Burundi from week 27 to week 37. A) Mpox-positive cases by age group; B) Total mpox-positive cases by age group and week of reporting; C) Tabular results of mpox-positive cases by age group. D) Median ages of mpox-positive cases by sex.

### Geographic distribution of mpox cases in Burundi

We assessed the geographic distribution of positive cases in Burundi (Figure 3). Mpox positive cases were identified from 14 provinces in Burundi with the majority of positive cases identified in Bujumbura Mairie (91/154; 59.1%) followed by Bubanza, Bujumbura, and Kayanza with 10 cases reported from each (6.5% of positive cases). Associations between geographic regions and age distribution of cases were limited due to the high proportion of overall mpox cases reported from Bujumbura Mairie and much lower proportions in any of the other provinces reporting. We did analyze age demographics in Bujumbura Mairie as compared to the complete national data set. When focusing on Bujumbura Mairie, where 59.1% (91/154) of all mpox cases were reported, children <15 years comprised 45% of cases (44/91), followed by those aged 15-30 years at 35.2% (32/91) and lastly those 30+ years with 16.5% of cases (15/91). When combining positive case data from all other provinces (63 cases total), children <15 years comprised the highest proportion of all cases at 65.1% (41/63), followed by those aged 15-30 years at 20.6% (13/63) and those 30+ years at 14.3% (9/63).

**Figure 3:**
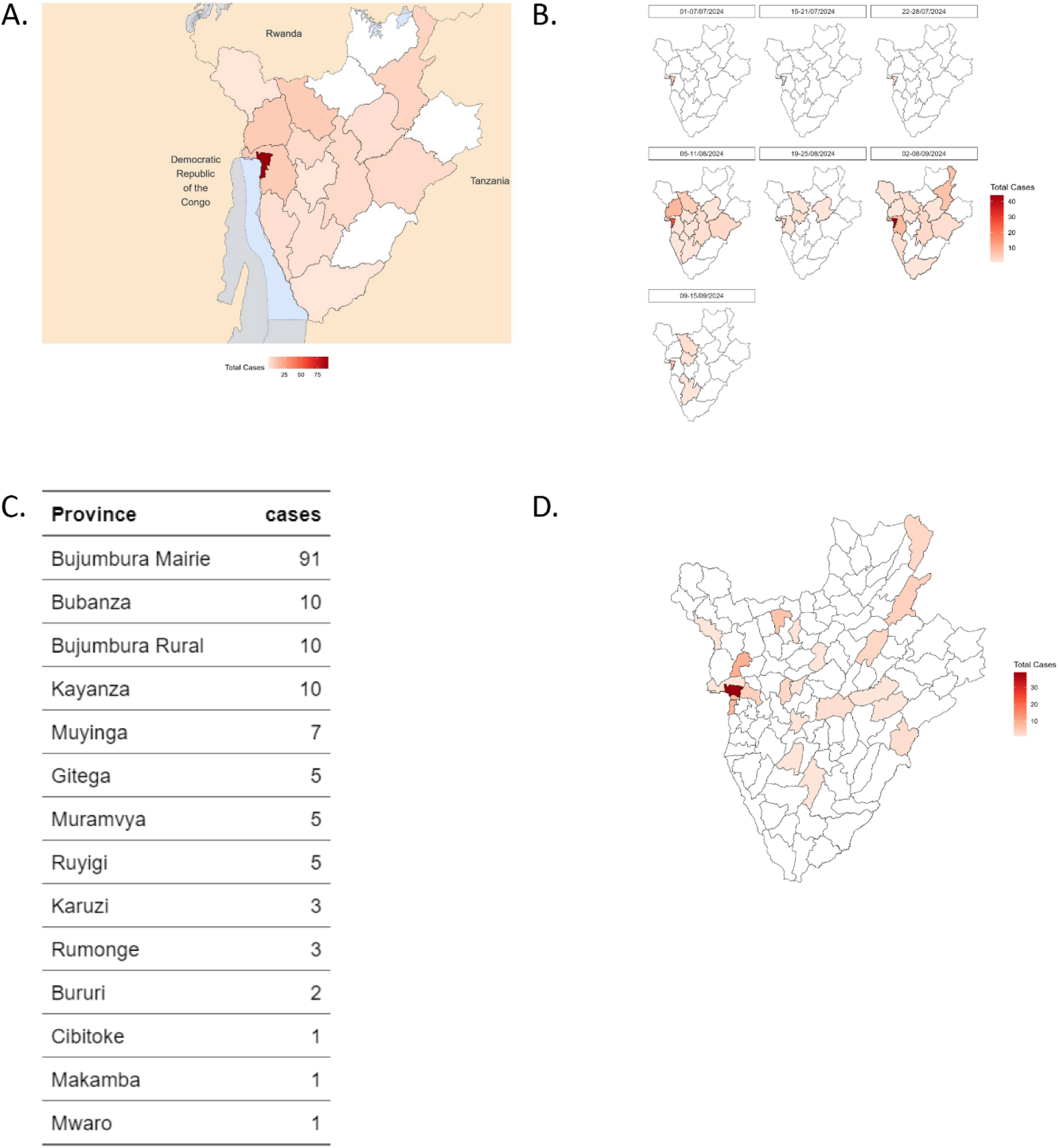
Mpox case mapping in Burundi from week 27 to week 37. A) Total mpox cases by province during reporting period. B) Total mpox cases by week across all provinces; C) Total cases by province (numerical); D) Total mpox cases by community during reporting period.

### Mpox-associated hospitalizations and clinical symptoms

Hospitalization data was available up to 25 September 2024 for confirmed mpox cases at the teaching hospital in Bujumbura and included total of 254 cases with 247 requiring care for more than one day (Table 1). The median age of those hospitalized was 19 (IQR: 8-28) with a median os 16 years (IQR: 7-25) and 22 (IQR: 8-32) for females and male patients, respectively. The overall duration of hospital admission was 8 days (IQR: 6-12) with 8 days (IQR: 5-9.8) and 9 days (IQR: 6-12) for females and males, respectively. No deaths were recorded though 45 patients remained hospitalized at the time of this report. The majority of patients has no known comorbidities. For hospitalized females, HIV positivity was noted among 6 patients, followed by hypertension (3 cases), asthma (2 cases), and diabetes (1 case). For hospitalized males, the most prominent comorbidities were hypertension (4 cases), asthma (3 cases), diabetes (3 cases), and HIV (1 case).

**Table 1:** Hospitalization data in Burundi up to 25 September 2024.

|  |  | Age (years) |  |  |
| --- | --- | --- | --- | --- |
|  | N | Median | IQR | p-value |
| **Males** | 132 | 22 | 8-32 | 0.013 |
| **Females** | 115 | 16 | 7-25 |  |
| **Total** | 247 | 19 | 8-28 |  |

We also analyzed clinical symptom and outcome data that was available for the hospitalized patients (250/254). Fever was reported for 50.4% of patients (126/250), with headache (19.6%; 49/250) and myalgia (1.2%; 3/250) reported less commonly. Lymphadenopathy was reported for 36.8% of patients (92/250). Rash was reported for nearly all patients (249/250) and with pustular or vesiculopapular rash most common (92%; 229/249). Generalized rash was most reported (78.3%; 195/249) and genital rash was reported among 18.1% of patients (49/249). Sore throat (including cough) was reported among 15.6% of patients (39/250). Nearly all hospitalized patients were reported as recovered or recovering (99.2%; 247/249); two cases of vision loss were reported among the outcomes (0.08%; 2/249).

## DISCUSSION

Here, we provide the first assessments of demographic, epidemiological, and hospitalization data from suspected and confirmed mpox cases in Burundi from 03 July 2024 to 09 September 2024. During this period, 607 suspected mpox cases underwent PCR testing with an overall mpox positivity rate of 25.4%. This report provides important insights into Clade Ib mpox cases. Sex-specific differences were present when considering age, mpox positivity, and hospitalization. While the median ages of males and females were very similar among mpox negative cases there was a large median age difference between males and females for mpox positive cases (17.5 years and 6 years, respectively), highlighting potential disparate sex-based epidemiologic risk factors for infection. Further case-investigation investigations are needed to better understand how exposures differ among males and females in Burundi and to better inform community recommendations on infection prevention and control

Among hospitalizations associated with mpox, there were no fatalities noted among these patients though there were individuals still hospitalized at the time of this reporting. These data provide important considerations for mpox response and containment efforts in Burundi. Among hospitalized patients, generalized rash was highly common among hospitalized patients with genital rash reported among 18.1% of patients with a median age of 23 years (IQR: 19-30). This suggests that the contributions of sexual (intimate) and non-sexual contact-mediated transmission needs further investigation in Burundi. Two reports of vision loss were included among the hospitalized patients with both cases being found in those >15 years. Notably, this early epidemiological data demonstrated that mpox cases during this period were overrepresented among children <15 years and the highest burdens being among those <9 years. Whether these data are reflective of certain activities or behaviours associated with greater risk of MPXV infection across sex and age groups needs to be addressed. An additional important consideration from this data is that while mpox cases in Burundi were overrepresented among children <15 years, the median age of hospitalization for both males and females was >15 years.

Considerations should include assessment of potential reporting biases by sex and age as well as activities among these groups that may be associated with higher infection risks (e.g. household caregiving, contact with contaminated materials). It is also prudent to consider recent data from North Kivu which included probable non-sexual (intimate) contact-based transmission events within and outside of households, including children (11). Thus, community engagement and messaging activities in Burundi should consider these early associations between mpox positivity status, sex, and age.

This report provides an early assessment of the demographic and epidemiological characteristics of Clade Ib mpox cases in Burundi and provide important insights for ongoing outbreak containment and mitigation strategies, including community engagement and messaging activities.

## ETHICS STATEMENT

Data was provided by country-wide mpox surveillance program. Permission to use this data was granted by the Institutional Ethics Committee at the Faculty of Medicine, University of Burundi (CIEB-FMCHUK no. 02/09/2024).

## DATA AVAILABILITY

All data produced in the present study are available upon reasonable request to the authors.

## REFERENCES

1. Breman JG, Kalisa R, Steniowski MV, Zanotto E, Gromyko AI, Arita I. Human monkeypox, 1970-79. Bull World Health Organ. 1980;58(2):165–82.

2. Foster SO, Brink EW, Hutchins DL, Pifer JM, Lourie B, Moser CR, et al. Human monkeypox. Bull World Health Organ. 1972;46(5):569–76.

3. Ladnyj ID, Ziegler P, Kima E. A human infection caused by monkeypox virus in Basankusu Territory, Democratic Republic of the Congo. Bull World Health Organ. 1972;46(5):593–7.

4. Van Dijck C, Hoff NA, Mbala-Kingebeni P, Low N, Cevik M, Rimoin AW, et al. Emergence of mpox in the post-smallpox era-a narrative review on mpox epidemiology. Clin Microbiol Infect. 2023;29(12):1487–92.

5. Okwor T, Mbala PK, Evans DH, Kindrachuk J. A contemporary review of clade-specific virological differences in monkeypox viruses. Clin Microbiol Infect. 2023;29(12):1502–7.

6. Ulaeto D, Agafonov A, Burchfield J, Carter L, Happi C, Jakob R, et al. New nomenclature for mpox (monkeypox) and monkeypox virus clades. Lancet Infect Dis. 2023;23(3):273–5.

7. Thornhill JP, Barkati S, Walmsley S, Rockstroh J, Antinori A, Harrison LB, et al. Monkeypox Virus Infection in Humans across 16 Countries - April-June 2022. N Engl J Med. 2022;387(8):679–91.

8. WHO. 2022-23 Mpox (Monkeypox) Outbreak: Global Trends. 2023.

9. Tarin-Vicente EJ, Alemany A, Agud-Dios M, Ubals M, Suner C, Anton A, et al. Clinical presentation and virological assessment of confirmed human monkeypox virus cases in Spain: a prospective observational cohort study. Lancet. 2022;400(10353):661–9.

10. Vakaniaki EH, Kacita C, Kinganda-Lusamaki E, O’Toole A, Wawina-Bokalanga T, Mukadi-Bamuleka D, et al. Sustained human outbreak of a new MPXV clade I lineage in eastern Democratic Republic of the Congo. Nat Med. 2024.

11. Mukadi-Bamuleka DK-LE, Mulopo-Mukanya N, Amuri-Aziza A, O’Toole A et al. First imported Cases of MPXV Clade Ib in Goma, Democratic Republic of the Congo: Implications for Global Surveillance and Transmission Dynamics. medRxiv. 2024.

